# Protocol for a scoping review of the impact of digital assistive technologies on the quality of life for people with dementia

**DOI:** 10.1101/2023.06.23.23291806

**Authors:** Charlotte Schneider, Tobias Kowatsch, Rasita Vinay

## Abstract

**Introduction:** Digital assistive technologies (e.g., applications, wearables, and robots) have emerged as promising tools for managing various aspects of daily life, such as basic assistance, encompassing social interaction, memory support, leisure activities, location tracking and health monitoring. In order to understand how these technologies can be utilized for people living with dementia, their impacts must first be reviewed. Currently, there is limited literature available on the topic, usually only focusing on a particular kind of digital assistive technology. Therefore, this paper presents a protocol for a scoping review that aims to provide a general overview of the impact digital assistive technologies can have on the quality of life for people living with dementia.

**Methods and analysis:** We will follow the scoping review framework proposed by Arksey and O’Malley. A comprehensive search will be performed to identify original research articles or clinical trials published between 2013 and 2023 across online databases. The review will encompass both qualitative and quantitative themes derived from the literature. Relevant studies will be identified through a comprehensive search using specific search terms related to the population (people with dementia), intervention (digital assistive technologies), and outcome (quality of life). The screening of titles, abstracts, and full texts will be performed to select eligible studies based on predetermined inclusion and exclusion criteria. Data will be extracted using a standardized form, and the findings will be synthesized and reported qualitatively and quantitatively.

**Ethics and dissemination:** Ethical approval is not required because this study is a scoping review based on published data. We intend to publish our findings in a peer-reviewed journal.

**OSF Registration:** https://osf.io/zcnx8/

**Strengths and limitations of this study:**

- This scoping review represents the first attempt to provide a comprehensive overview of digital assistive technologies and their impact on quality of life for PWD
- An extensive search strategy will be implemented, covering five electronic databases, spanning a period of ten years.
- However, given the rapidly expanding field of digital health technologies, it is possible that this systematic review may overlook ongoing or planned studies.

## Introduction

In 2023, more than 55 million individuals worldwide are affected by dementia, with approximately 10 million new cases reported every year [1]. Over 60% of this population live in low- and middle-income countries [1]. Dementia encompasses various impairments regarding memory, cognition, and the ability to perform daily activities [1]. It progressively worsens over time and primarily affects older individuals (over the age of 65), although not everyone will experience it. There are also possibilities for individuals younger than 65 years of age to develop dementia, known as young onset dementia. Globally, dementia currently ranks as the seventh leading cause of death, significantly contributing to disability and dependency among the older population [1]. This demographic shift poses challenges for caregivers and our healthcare system, prompting increased attention towards mitigating these burdens through digital assistive technologies to sustain the independence of people with dementia (PWD) [2].

Digital assistive technologies can help individuals and caregivers manage aspects of their daily lives. They are promising tools for the care and support of elderly people and also help to ease the burdens of caregiving. Advancements in technology have led to the development of devices and applications that use sensory data specifically for PWD. For instance, smartphones and wearables are being utilized to monitor physical activities, enabling homecare assistance [3], or as location trackers to monitor wandering behavior [4]. Furthermore, with the rise of Artificial Intelligence (AI), there have been developments of Smart Assistive Robots (SAR), which can assist PWD by providing companionship and engaging in pet therapy, for example, as demonstrated by the robotic seal Paro [5]. These technologies go beyond mere assistance in daily activities, as they also aid in maintaining social interaction, memory support, participation in leisure activities, location tracking, and health monitoring [2][6].

To this end, maintaining a good quality of life is essential for PWD and must be considered when assessing the impact of digital assistive technologies. Quality of life encompasses physical and mental health, as well as social and emotional well-being (e.g., emotional stability, social integration, or self-esteem) [7]. Quality of life can be measured with different instruments, such as questionnaires and self-rating scales for the individual’s overall perceived quality of life, and also through activity instruments or cognitive status assessments [8]. These measures and instruments must therefore also be considered as guiding tools for determining the quality of life for a PWD.

This scoping review aims to provide a general overview of the impact digital assistive technologies can have on the quality of life for PWD, due to the lack of existing literature reviews on the topic. Through this scoping review, we hope to explore the opportunities and potential benefits that digital technology can offer in improving caregiving and living standards of PWD.

Furthermore, this review serves as a tool to create greater awareness among various stakeholders, including policymakers, researchers, politicians, and even management teams of elderly care companies and institutions. By presenting a synthesis of current evidence, it can strengthen the decision-making process by enabling stakeholders to understand what digital assistive technologies are available and what works effectively in enhancing the quality of life as a goal of care.

## Methodology

### Scoping review

We will use the framework proposed by Arksey and O’Malley for this review [9]. Therefore, the scoping review will follow the following five-step process: (i) identifying the research question, (ii) identifying relevant studies, (iii) selecting eligible studies, (iv) charting the data, and (v) collating and summarizing the results. We will follow the *Preferred Reporting Items for Systematic Reviews and Meta-Analyses* (PRISMA) Protocols [10]. As the main aim of this scoping review is to describe the state of the literature, a quality assessment will not be conducted as generally done for a systematic review.

This scoping review has been preregistered on OSF Registries (https://osf.io/zcnx8/).

### Identifying the research question

The main research question this review aims to answer is, “what is the impact of digital assistive technologies on the quality of life for people with dementia?”. The findings will present both qualitative and quantitative themes surrounding the research question, providing a current overview of the impact of digital assistive technologies on the quality of life for people with dementia, as reflected in the literature. The research question was formulated using the PIO concept (see Table **1**) according to prior work [11].

**Table 1.** The PIO framework for the eligibility of studies.

| Concept | Determinants |
| --- | --- |
| P - Population | People with dementia |
| I - Intervention | Digital assistive technologies |
| O - Outcome | Quality of life |

### Identifying relevant studies

The search terms and strategy used in this scoping review are summarized in Table 2 for each PIO concept. Search terms were derived from a preliminary search and analyzed by comparing the words found in titles, abstracts, and keywords. Additionally, to enhance the accuracy and comprehensiveness of the search results, all authors were involved in a consensus process, and an additional expert was consulted to validate the identified terms and suggest any additional relevant keywords.

**Table 2.**
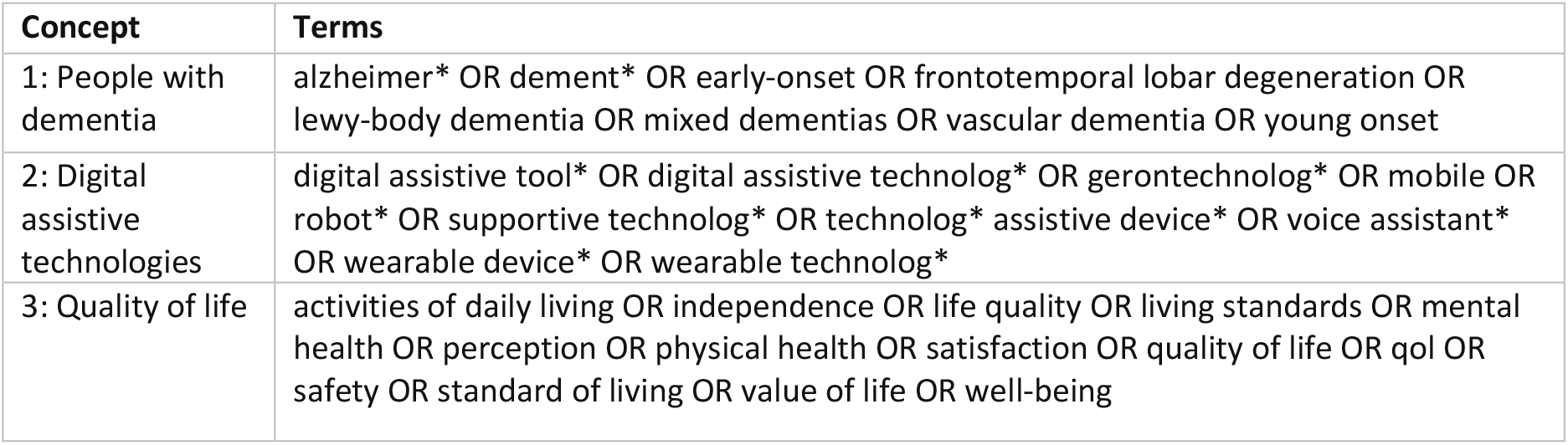
The search terms derived for the PIO framework.

| Concept | Terms |
| --- | --- |
| 1: People with dementia | alzheimer* OR dement* OR early-onset OR frontotemporal lobar degeneration OR lewy-body dementia OR mixed dementias OR vascular dementia OR young onset |
| 2: Digital assistive technologies | digital assistive tool* OR digital assistive technolog* OR gerontechnolog* OR mobile OR robot* OR supportive technolog* OR technolog* assistive device* OR voice assistant* OR wearable device* OR wearable technolog* |
| 3: Quality of life | activities of daily living OR independence OR life quality OR living standards OR mental health OR perception OR physical health OR satisfaction OR quality of life OR qol OR safety OR standard of living OR value of life OR well-being |

A comprehensive search will be performed across five electronic databases: Cochrane, Embase, PubMed, Scopus, and Web of Science, to locate published literature surrounding the research question. The search strategy exclusively considers articles published between 2013 and 2023 to focus on recent technological advancement, allowing for a more up-to-date review.

The terms from Table 2 are applied to each database, scanning for the title, abstract, and, if available, Medical Subject Headings (MeSH) terms; otherwise, keywords. The Boolean operator OR is utilized within each concept, and each concept is then linked together using the AND operator.

### Selection of eligible studies

The screening of titles and abstracts will be guided by the PIO framework (see Table **1**), following the eligibility criteria in Table **3** to ensure the relevance of the included studies to the research question.

**Table 3.** Inclusion and Exclusion Criteria.

| Inclusion Criteria | Exclusion Criteria |
| --- | --- |
| • Original Research Articles or Clinical Trials (completed)<br>• Articles which have a primary focus on digital assistive technologies for PWD<br>• Articles discussing perspectives of caregivers, family members, or healthcare workers in relation to a PWD<br>• Articles about people living in diverse settings including communities, hospitals or nursing homes, and all severities of dementia. We will not use age as a criterion. | • Articles not in English or German<br>• Articles which discuss dementia negligibly or with other comorbidities/ health conditions<br>• Articles that mention digital assistive technologies briefly or as an insignificant part of a review<br>• Pilot or feasibility studies which only report the implementation of an intervention |
|  | • Book chapters, commentaries, conference proceedings, editorials, interviews, opinion pieces, proposals, reports, protocols, short news<br>• Non-human studies |

On May 17, 2023, a literature search was conducted across the electronic databases previously mentioned, resulting in 5’027 articles and trials (see supplementary file 1). The search results were extracted and uploaded onto a literature review software, Rayyan (https://www.rayyan.ai) for screening. From May to July 2023, authors CS and RV intend to conduct title and abstract screening of all eligible articles to determine their suitability for a full-text review according to the inclusion and exclusion criteria. Author TK will be involved if substantial discrepancies cannot be resolved through discussion and consensus. The level of agreement between the reviewers will be calculated and reported. To ensure reliability, authors CS and RV will conduct a full-text review to determine their inclusion in the study. Author CS will critically analyze the final sample of studies, and all authors will be involved in the charting process.

The selection process will follow the recommendations in the *Preferred Reporting Items for Systematic Reviews and Meta-Analyses Extension for Scoping Reviews (PRISMA-ScR) checklist* [12] and be mapped using the PRISMA-P chart [10].

### Charting the data

A data extraction form will be used to capture relevant information from each included article. Charting the data is planned for July 2023. An example of the data extraction form is described in Table **4**.

**Table 4.** Data Charting Form.

|  |  |
| --- | --- |
| Title of study |  |
| DOI |  |
| Year of publication |  |
| Author name/s |  |
| Author location/s |  |
| Study approach | i.e., qualitative or quantitative or mixed method |
| Type of article | i.e., Case Study, Observational Study, RCT, Review, Trial, Other... |
| Study location | i.e., where the study was carried out |
| Class of digital assistive technology | i.e., AI, Application, AR/ VR, Conversational Agent, Wearable, Other... |
| Explanation of the digital assistive technology | i.e., specification of the digital assistive technology (e.g. name / brand) |
| Sensory distribution channel | i.e., acoustic, proprioceptive, tactile, visual |
| Target population |  |
| Outcome measured | i.e., the primary outcome being measured in the study |
| Aim/s of the study |  |
| Study methods summary |  |
| Key findings | i.e., study findings relevant to study objectives |
| Quality of life measure | i.e. how is quality of life measured (e.g. rating of Quality of life through an questionnaire, activity instrument, cognitive status, etc.) |
| Reported effect |  |
| Notes |  |

### Collating, summarizing, and reporting the results

After conducting full-text reviews using Table **4**, a final list of studies for the scoping review will be constructed in August. These articles will be critically analyzed, and the main findings will be reported. Data will be analyzed both qualitatively and quantitatively. Quantitative analysis will include (1) author locations, (2) study approach (3) type of article (4) study locations, (5) class of digital assistive technology, (6) sensory distribution channel (7) target population, (8) outcome being measured, and (9) instrument measuring quality of life. From a pilot study of the articles, it can be seen that there is rarely a specific “quality of life instrument” is used. Indirect outcomes, which also influence the quality of life, are therefore recorded (e.g., activity instruments, cognitive status, rating of the patient’s quality of life[8]). Qualitative analysis will help identify the impact of the digital assistive interventions on the target population in each study.

## Discussion

The proposed scoping review aims to demonstrate the impact digital assistive technologies can have on the quality of life for PWD. Through this review, we hope to create greater awareness of the different digital assistive technologies that have been researched, not only for PWD, but also their carers. Ultimately, the outcomes of this review will provide evidence-based insights to health policymakers and stakeholders, enabling them to address the pressing needs of an increasingly affected population. The findings will contribute to shaping policies, resource allocation, and interventions that effectively leverage digital technologies to improve the quality of care and support available to PWD and their caregivers.

A limitation of this review is that certain digital technologies may be missing due to the search terms selected, as there is no uniform definition of “digital assistive technologies”. Another limitation is the lack of a market analysis to provide an outlook of companies which already provide digital assistive technologies to individuals with a cognitive impairment – such as those with dementia, and is therefore recommended for future research. Moreover, due to the absence of a quality appraisal, it is not possible to make any remarks regarding the reliability of the study interventions on the measured outcome. A risk of bias of the evidence or methodological limitations was also not assessed, given the focus of the scoping review.

## Data Availability

All data produced in the present study are available upon reasonable request to the authors

## Acknowledgment

The authors would like to thank Dr. med. Mathias Schlögl for his valuable input and feedback throughout the process.

## Author contributions

All authors have made substantial intellectual contributions to the development of this protocol and its revisions. The search question was conceptualized by TK and RV, and further developed by CS. The review approach and design was conceptualized by RV, with advice from TK. CS and RV developed and tested search terms with input and revisions from TK. CS initiated drafting of the manuscript followed by further iterations after substantial input and appraisal from TK and RV. All authors approved the final version of the manuscript.

